# GLP-1 Receptor Agonist Initiation Is Associated With Excess Skeletal Muscle Loss in Adults With Diabetes: A Prospective Population-Based MRI Study

**DOI:** 10.64898/2026.09.15.26363157

**Authors:** Era Stambollxhiu, Jannik Luebberstedt, Miriam Kumpf, Stefan N. Willich, Thomas Keil, Tobias Pischon, Thoralf Niendorf, Annette Peters, Sebastian Ziegelmayer, Marcus R. Makowski, Keno K. Bressem, Lisa C. Adams

## Abstract

**Objective:** To determine whether glucagon-like peptide 1 receptor agonist (GLP-1 RA) initiation is associated with excess skeletal muscle loss on repeat whole-body MRI.

**Research Design and Methods:** In the German National Cohort, 58 adults with diabetes who initiated a GLP-1 RA between whole-body MRI examinations 4.6 years apart were matched on sex, age, BMI, and interval to 130 never-exposed controls with diabetes. Deep-learning-segmented tissue volumes were compared within matched sets.

**Results:** Skeletal muscle volume declined 2.75 percentage points more in initiators (95% CI -4.51 to -0.99; P = 0.003), with 2.54 percentage points greater weight loss (P = 0.021). Adipose tissue differences (−4.8% internal torso, -3.2% subcutaneous) were nonsignificant.

**Conclusions:** In this first population-based comparison of whole-body MRI before and after GLP-1 RA initiation with matched unexposed controls, initiators lost roughly 60% more muscle. Skeletal muscle loss is a measurable component of GLP-1 RA weight loss in routine care.

**Article Highlights:**

- **Why did we undertake this study?** Rapid uptake of GLP-1 RAs has outpaced evidence on long-term muscle change during routine treatment.
- **What is the specific question we wanted to answer?** We asked whether GLP-1 RA initiation was associated with excess muscle and fat loss versus matched controls with diabetes.
- **What did we find?** Over 4.6 years, initiators lost roughly 60% more skeletal muscle than matched controls (−2.75 percentage points) alongside greater weight loss. Each additional kilogram lost was accompanied by about 100 mL of additional muscle loss.
- **What are the implications of our findings?** Skeletal muscle loss is a measurable component of GLP-1 RA-associated weight loss in routine care. Muscle mass should be monitored during treatment, and muscle-preservation strategies such as resistance training and adequate protein intake need evaluation.

---

Glucagon-like peptide 1 receptor agonists (GLP-1 RAs) are transforming glucose and weight management in type 2 diabetes, yet the quality of the weight lost during routine treatment remains an evidence gap. Meta-analyses of randomized trials suggest that approximately one quarter to one third of weight loss with incretin-based therapy is lean mass (1–3). Lean mass is not equivalent to skeletal muscle, and longitudinal imaging evidence remains limited. Because type 2 diabetes itself accelerates muscle loss (4), any excess attributable to treatment must be judged against contemporaneous controls with diabetes. To address this gap, we used repeat whole-body MRI from the population-based German National Cohort (NAKO) to test whether adults with diabetes who initiated a GLP-1 RA experienced excess skeletal muscle and adipose tissue loss compared with matched adults with diabetes who remained unexposed.

## Research Design and Methods

NAKO is a population-based prospective cohort that recruited 205,415 participants across Germany between 2014 and 2019. Approximately 30,000 underwent standardized whole-body MRI on 3-T scanners (Magnetom Skyra, Siemens Healthineers, Erlangen, Germany), with repeat imaging after approximately 4-5 years (5,6). All participants provided written informed consent, and NAKO was approved by the ethics committees of the participating study centers. This analysis was conducted under NAKO use-and-access application no. 1004.

Medication use was recorded at both examinations in standardized interviews. Initiators reported no GLP-1 RA use at baseline (U1) and use at follow-up (U2). Controls reported no use at either examination, and participants using a GLP-1 RA at U1 were excluded. The specific agent, initiation date, dose, and adherence were unavailable. Body composition was quantified from the whole-body T1-weighted Dixon sequence, using TotalSegmentator MRI (version 2.15.0), an automated deep-learning segmentation tool (7). All segmentation masks were visually reviewed by a resident radiologist with 2 years of experience (E.S.) and a board-certified radiologist with more than 10 years of experience (L.C.A.). Only two masks were manually corrected and none of the segmentations had to be excluded for segmentation failure or incomplete coverage. Outcomes were volumes of skeletal muscle, subcutaneous adipose tissue, and internal non-subcutaneous torso adipose tissue, the last comprising visceral and other deep trunk depots.

Diabetes was defined by self-reported diagnosis, HbA1c ≥6.5% (48 mmol/mol), or glucose-lowering medication use at U1. Each initiator was matched without replacement to up to three never-exposed controls with diabetes, using exact matching on sex and calipers of 3 years for age, 2 kg/m² for BMI, and 1 year for the interval between examinations. When more than three controls were eligible, the three closest by summed standardized squared distance were selected. Initiators without an eligible control were not matched.

For each matched set, the initiator’s change from U1 to U2 was compared with the mean change of the matched controls, yielding one contrast per set. Percentage change was calculated as 100 × (U2 - U1)/U1. Muscle and weight effects are therefore reported as between-group differences in percentage points. A one-sample t test of the set-level contrasts provided estimates, 95% CIs, and two-sided P values. Skeletal muscle volume was the primary outcome. Internal torso adipose tissue was analyzed as a log ratio and back-transformed, and subcutaneous adipose tissue was analyzed in milliliters. A linear regression related the set-level muscle-volume contrast in milliliters to the weight contrast in kilograms. The slope quantifies muscle loss per additional kilogram of weight loss, and the intercept quantifies the excess muscle loss at equal weight change. Analyses were performed in Python version 3.14.2 (Python Software Foundation) using pandas 3.0.1, NumPy 2.4.2, SciPy 1.18.0, and statsmodels 0.14.6.

## Data and Resource Availability

NAKO data are available to researchers through the formal use-and-access process (https://nako.de/). Data for this analysis were obtained under application no. 1004.

## Results

Of 65 initiators, 63 had whole-body Dixon MRI at both examinations and 62 had a measured baseline BMI. Fifty-nine were matched to controls. One was subsequently excluded because no matched control had usable imaging, leaving 58 matched sets comprising 58 initiators and 130 controls (Fig. 1). Of these, 25 sets had three controls, 22 had two, and 11 had one. The interval between examinations was approximately 4.6 years. Initiators and controls were similar in age, sex, BMI, and overall use of glucose lowering medication including insulin (absolute standardized mean differences ≤0.12), whereas initiators were taller and heavier, had higher blood pressure and HbA1c, and less often reported a diabetes diagnosis, and more often used SGLT2 inhibitors and sulfonylureas but less often metformin. Seven initiators and 22 controls met the diabetes definition solely by HbA1c ≥6.5% (48 mmol/mol) (Table 1).

**Figure 1.**
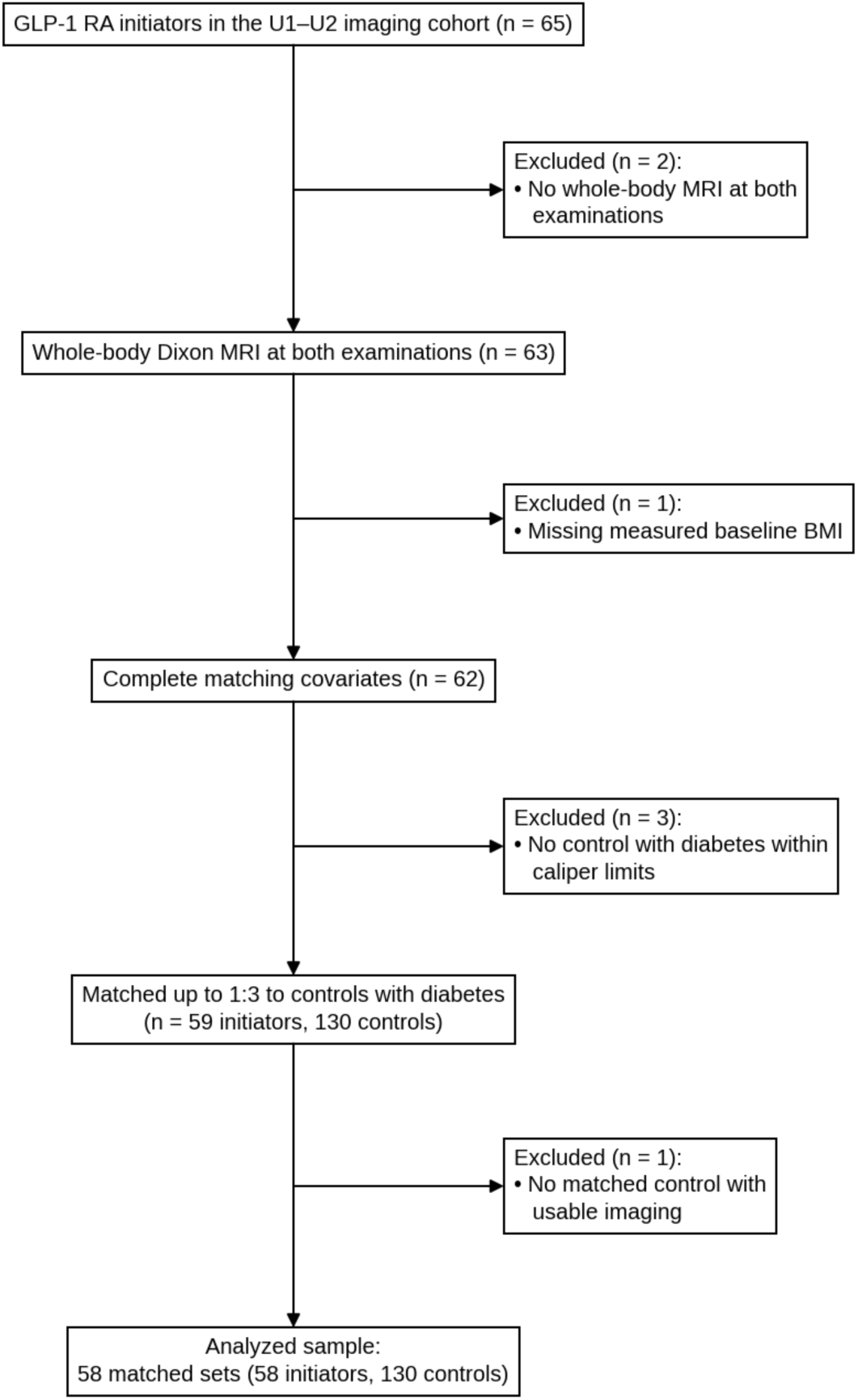
Cohort flow to the analyzed sample. Of 65 adults with diabetes who reported GLP-1 RA initiation between examinations, 58 initiators and 130 matched controls with diabetes entered the analysis. Each initiator was matched to up to three controls. Boxes on the main path show participants retained, and side boxes show exclusions.

**Table 1.** Baseline (U1) characteristics of GLP-1 RA initiators and their matched controls with diabetes (58 matched sets).

| Characteristic | GLP-1 RA initiators (n = 58) | Matched controls with diabetes (n = 130) | SMD |
| --- | --- | --- | --- |
| Age (years) | 55.9 (7.5) | 56.8 (7.2) | -0.12 |
| Female | 17 (29%) | 41 (32%) | -0.05 |
| BMI (kg/m<sup>2</sup>) | 33.5 (4.6) | 33.0 (4.3) | 0.11 |
| Weight (kg) | 102.8 (17.8) | 98.2 (17.0) | 0.26 |
| Height (cm) | 174.9 (9.4) | 172.1 (9.3) | 0.30 |
| Self-reported diabetes diagnosis | 41 (71%) | 106 (82%) | -0.26 |
| Age at diabetes diagnosis (years) | 49.0 (8.8) | 47.9 (11.6) | 0.10 |
| HbA1c (%) | 7.4 (1.7) | 7.0 (1.2) | 0.27 |
| HbA1c (mmol/mol) | 57 (19) | 53 (13) | 0.27 |
| Biguanides (Metformin) use | 19 (33%) | 59 (45%) | -0.26 |
| Insulin use | 11 (19%) | 21 (16%) | 0.07 |
| SGLT2 inhibitor use | 7 (12%) | 7 (5.4%) | 0.24 |
| Sulfonylurea use | 4 (6.9%) | 4 (3.1%) | 0.18 |
| Hypertension | 29 (50%) | 48 (37%) | 0.27 |
| Systolic BP (mmHg) | 138.1 (14.4) | 133.9 (15.6) | 0.28 |
| Diastolic BP (mmHg) | 84.0 (10.1) | 80.6 (9.8) | 0.34 |
| Antihypertensive use | 40 (69%) | 82 (63%) | 0.12 |
| β-Blocker use | 20 (34%) | 39 (30%) | 0.10 |
| Lipid-lowering use | 19 (33%) | 45 (35%) | -0.04 |
| Statin use | 18 (31%) | 42 (32%) | -0.03 |
| Physical activity (MET-min/week) | 2400 (560, 9780) | 3680 (720, 10080) | -0.11 |

Data are mean (SD) unless otherwise indicated. Physical activity is median (25th, 75th percentile).
Categorical variables are n (%). Glucose-lowering medication classes are not mutually exclusive.
BMI, body mass index; BP, blood pressure; GLP-1 RA, glucagon-like peptide 1 receptor agonist;
HbA1c, glycated hemoglobin; MET, metabolic equivalent of task; SGLT2, sodium-glucose
cotransporter 2; SMD, standardized mean difference (initiators minus controls); U1, baseline
examination.

Mean skeletal muscle volume declined by approximately 7.1% in initiators and 4.5% in controls, a roughly 60% greater loss in initiators (Table 2). The matched-set difference was -2.75 percentage points (95% CI -4.51 to -0.99; P = 0.003), corresponding to -573 mL (95% CI -936 to -210). Adjusting for baseline HbA1c did not alter the muscle contrast (Supplementary Table 1). Body weight also declined more in initiators (difference -2.54 percentage points, 95% CI -4.69 to -0.40; P = 0.021). Internal torso adipose tissue showed a ratio of 0.952 (95% CI 0.894-1.015; P = 0.128), corresponding to an estimated 4.8% greater decline, and the subcutaneous adipose tissue difference was -792 mL (95% CI -1,868 to 284; P = 0.146), approximately 3.2% of mean baseline volume (Supplementary Fig. 2). Each additional kilogram of weight loss relative to controls was associated with 101 mL (95% CI 76-127) of additional skeletal muscle volume loss, so approximately 10% of each additional kilogram lost was skeletal muscle. At equal weight change, initiators lost 267 mL (95% CI -14 to 547) more muscle than controls (regression intercept, R² = 0.47). Individual weight responses varied widely and overlapped between groups (Supplementary Fig. 1). Representative segmentations are shown in Fig. 2.

**Figure 2.**
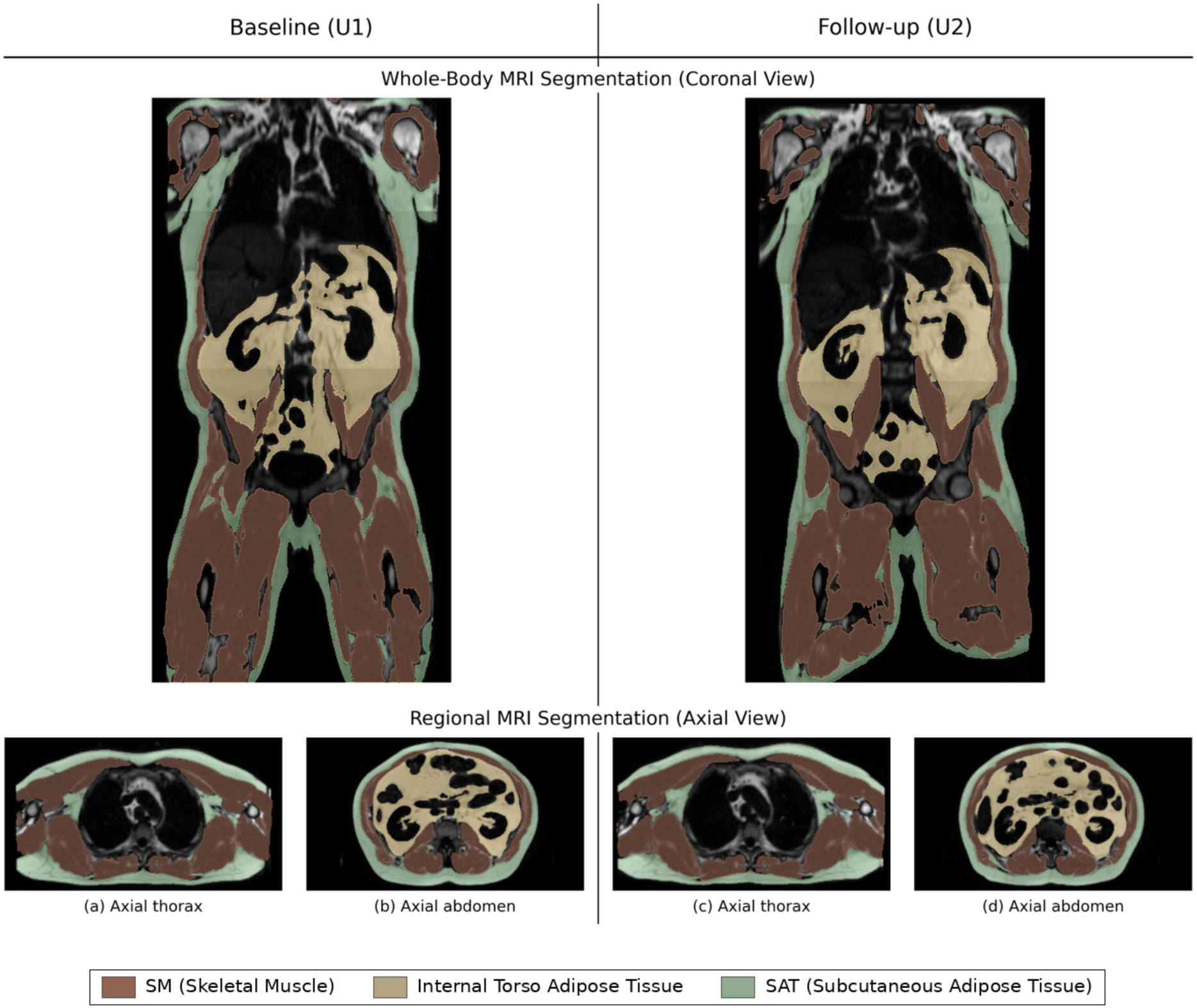
Automated segmentation of whole-body Dixon MRI in one GLP-1 RA initiator at baseline (U1) and follow-up (U2). Coronal whole-body and regional axial views show skeletal muscle in brown, internal torso adipose tissue (visceral and other deep trunk depots) in yellow, and subcutaneous adipose tissue in green. The images are illustrative and are not used to infer an individual treatment effect.

**Table 2.** Skeletal muscle and adipose tissue volumes and body weight at baseline (U1) and follow-up (U2) in GLP-1 RA initiators and matched controls with diabetes, with matched-set contrasts (58 matched sets).

| Outcome | GLP-1 RA initiators (n = 58) |  |  | Matched controls with diabetes (n = 130) |  |  | Matched-set contrast (95% CI) | P |
| --- | --- | --- | --- | --- | --- | --- | --- | --- |
|  | U1 | U2 | Change (%) | U1 | U2 | Change (%) |  |  |
| Skeletal muscle volume (mL) | 19,835<br>(3,518) | 18,442<br>(3,482) | -7.1<br>(5.0) | 19,034<br>(3,541) | 18,188<br>(3,579) | -4.5<br>(5.7) | -2.75 pp (-4.51 to -0.99) | 0.003 |
| Body weight (kg) | 102.8<br>(17.8) | 98.0<br>(17.5) | -4.6<br>(6.1) | 98.2<br>(17.0) | 96.6<br>(17.7) | -1.6<br>(6.7) | -2.54 pp (-4.69 to -0.40) | 0.021 |
| Internal torso adipose tissue (mL)<sup>‡</sup> | 7,007<br>(2,008) | 6,846<br>(2,061) | -0.9<br>(16.7) | 6,277<br>(2,310) | 6,440<br>(2,409) | +3.9<br>(19.5) | Ratio 0.952 (0.894 to 1.015) | 0.128 |
| Subcutaneous adipose tissue (mL) | 24,775<br>(8,089) | 23,350<br>(7,900) | -5.4<br>(11.6) | 24,784<br>(7,545) | 24,205<br>(7,558) | -1.9<br>(11.7) | -792 mL (-1,868 to 284) | 0.146 |

Data are mean (SD); n = 58 initiators and 130 controls for every cell. Change (%) is the mean of within-person percentage changes, $100 \times (U2 - U1)/U1$ , and therefore differs slightly from the change between the group means shown. Matched-set contrasts are within-set differences (initiator minus mean of matched controls) from one-sample t tests. <sup>‡</sup>Internal torso adipose tissue was analyzed as a log ratio; the change shown is the geometric mean change, back-transformed from the mean within-person log ratio (SD of the log ratio 0.18 in both groups). GLP-1 RA, glucagon-like peptide 1 receptor agonist; pp, percentage points; U1, baseline examination; U2, follow-up examination.

## Conclusions

In this matched population-based imaging cohort, GLP-1 RA initiators lost roughly 60% more skeletal muscle over 4.6 years than never-exposed controls with diabetes, an excess of 2.75 percentage points that corresponds to almost three additional years of age-related decline at the rate observed in controls. To our knowledge, this is the first population-based study to combine standardized whole-body MRI before and after GLP-1 RA initiation with a matched unexposed comparison group. Because both groups had diabetes and were followed contemporaneously, this separation isolates the muscle loss associated with GLP-1 RA initiation from that of diabetes and aging.

Previous longitudinal imaging studies have reported within-person body-composition changes after semaglutide or regional muscle changes among GLP-1 RA users, while the SURPASS-3 MRI substudy found that thigh muscle volume loss with tirzepatide broadly tracked the accompanying weight change (8–10). Our whole-body analysis extends this evidence beyond trial-derived lean mass and regional muscle measures and adds a matched unexposed comparison. Whether muscle loss with GLP-1 RAs is an adaptive consequence of weight reduction or exceeds it remains debated (11). In our data, approximately 10% of each additional kilogram lost was skeletal muscle, consistent with muscle loss accompanying weight reduction generally (1,2,12), and the residual excess at equal weight change did not reach statistical significance. Relative adipose tissue losses were of similar or larger magnitude but imprecisely estimated, so initiators lost muscle alongside fat.

This analysis has limitations. The specific agent, dose, and time since initiation were not available for the follow-up examination, so exposure duration varied within the 4.6-year window, and some controls may have used a GLP-1 RA transiently between examinations. Both misclassifications bias toward the null and make our estimate conservative. The findings characterize GLP-1 RAs as a class in the diabetes indication and should not be extrapolated to dual agonists or to obesity treatment without diabetes. Although matching balanced age, sex, BMI, and follow-up interval, and overall glucose-lowering medication use was similar, initiators had higher baseline HbA1c and more intensified therapy (Table 1). Adjustment for baseline HbA1c did not materially change the muscle estimate (Supplementary Table 1), although residual confounding by indication cannot be excluded. Given the scale of GLP-1 RA use, monitoring of muscle mass and muscle-preservation strategies such as resistance training and adequate protein intake deserve prospective evaluation alongside these therapies, including effects on muscle strength and function. This applies particularly to older adults with diabetes, who are already at increased risk of sarcopenia (4,13,14).

## Data Availability

This project was carried out with data (application number NAKO-1004) from NAKO. NAKO is funded by the Federal Ministry of Research, Technology and Space (01ER1301A/B/C, 01ER1511D, 01ER1801A/B/C/D, and 01ER2301A/B/C), the federal states of Germany and the Helmholtz Association, the participating universities, and the institutes of the Leibniz Association. We thank all participants who took part in the NAKO study and the staff of this research initiative.

## Author Contributions

E.S.: Conceptualization, Methodology, Software, Validation, Formal analysis, Investigation, Data curation, Writing - Original draft, Visualization; J.L.: Conceptualization, Methodology, Software, Validation, Formal analysis, Investigation, Data curation, Writing - Original draft, Visualization; M.K.: Writing - Review & Editing, Visualization; S.N.W.: Writing - Review & Editing; T.K.: Writing - Review & Editing; T.P.: Writing - Review & Editing; T.N.: Writing - Review & Editing; A.P.: Writing - Review & Editing; S.Z.: Writing - Review & Editing; M.R.M.: Writing - Review & Editing; K.K.B.: Conceptualization, Methodology, Software, Investigation, Resources, Writing - Original draft, Supervision, Project administration; L.C.A.: Conceptualization, Methodology, Software, Investigation, Resources, Writing - Original draft, Supervision, Project administration. All authors reviewed and edited the manuscript and approved the final version. E.S. is the guarantor of this work and, as such, had full access to all the data in the study and takes responsibility for the integrity of the data and the accuracy of the data analysis.

