## Supplementary Material for "GLP-1 Receptor Agonist Initiation Is Associated With Excess Skeletal Muscle Loss in Adults With Diabetes: A Prospective Population-Based MRI Study"

Online-only supplemental material. Contents: Supplementary Fig. 1, Supplementary Fig. 2, Supplementary Table 1

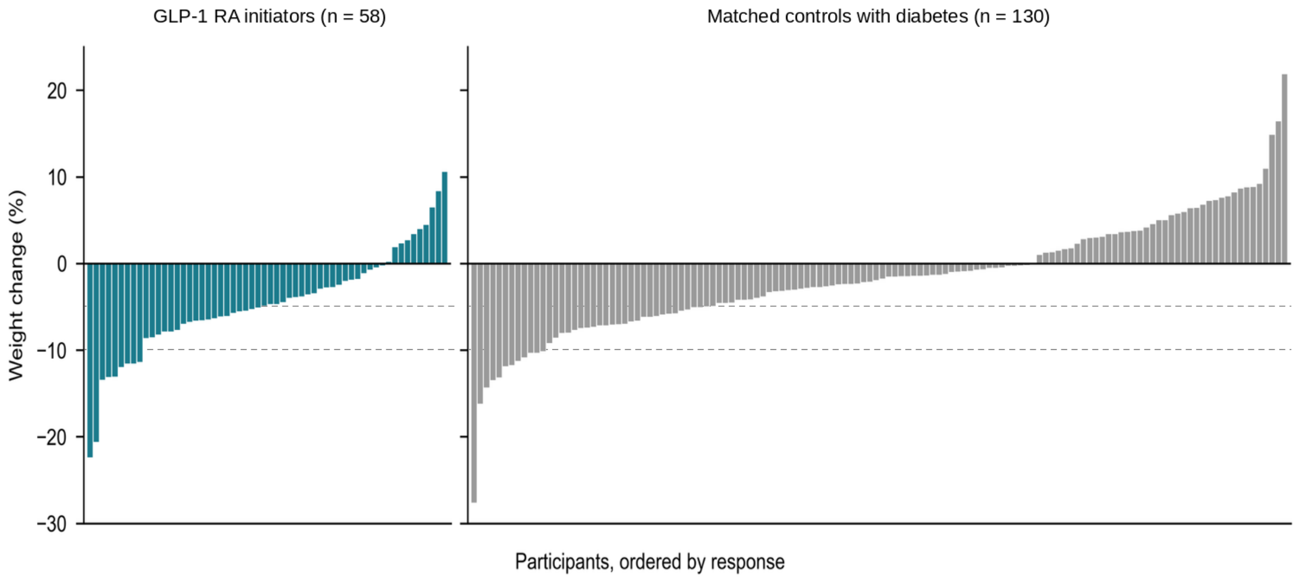


### Supplementary Figure 1

**Supplementary Fig. 1.** Individual percentage weight change from baseline to follow-up, one bar per person, for GLP-1 RA initiators and matched controls with diabetes. Each initiator had up to three controls, so the panels differ in size. Dashed lines mark -5% and -10%. The formal estimate used matched-set contrasts rather than the marginal distributions. (Online-only supplemental material)

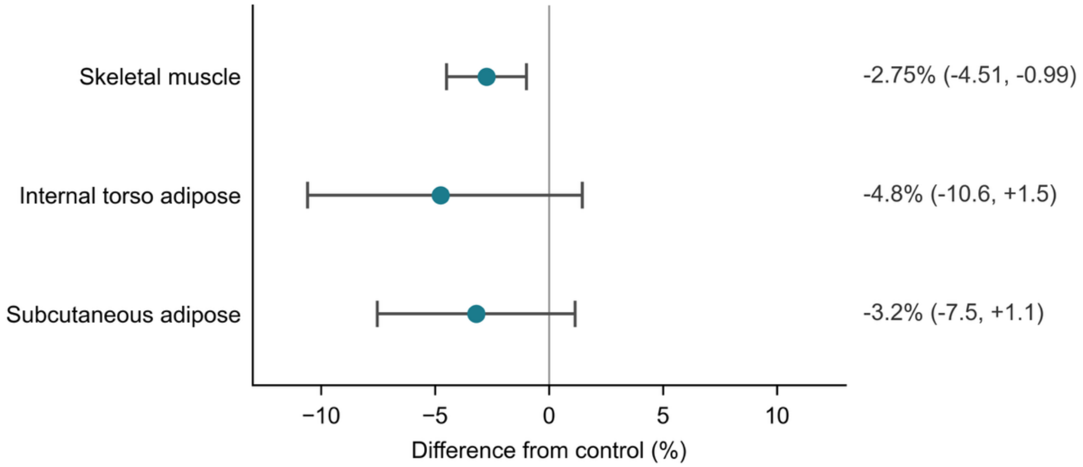


### Supplementary Figure 2

**Supplementary Fig. 2.** Matched-set differences in tissue-volume change from baseline to follow-up among 58 GLP-1 RA initiators and their matched controls with diabetes. Points represent mean within-set contrasts (initiators minus controls), and error bars represent 95% CIs. For visual comparison only, all estimates are displayed on a common percentage scale. Skeletal muscle is shown as the between-group difference in percentage change, internal torso adipose tissue as 100 × (ratio - 1) derived from the back-transformed mean log ratio, and subcutaneous adipose tissue as the absolute volume difference divided by the mean baseline volume of the matched analysis sample. Negative values indicate a greater decline in initiators. Numerical estimates on the prespecified analysis scales are reported in Results. (Online-only supplemental material)

### Supplementary Table 1

**Supplementary Table 1.** Set-level ordinary least squares regression of the primary muscle contrast (initiator percentage change minus the mean of matched controls) on the initiator-minus-control difference in baseline HbA1c, centered, so that the intercept is the contrast at equal HbA1c. Three initiators lacked HbA1c and their sets were excluded from the adjusted models. Control means used the available controls.

| **Model** | Matched sets (n) | Contrast (percentage points) | 95% CI | P |
|---|---|---|---|---|
| 1. Unadjusted (full sample) | 58 | -2.75 | -4.51 to -0.99 | 0.003 |
| 1b. Unadjusted (HbA1c-complete sets) | 55 | -2.9 | -4.73 to -1.07 | 0.002 |
| 2. HbA1c-adjusted (intercept = contrast at equal HbA1c) | 55 | -2.9 | -4.74 to -1.07 | 0.003 |
